# Modelling the association between neutralizing antibody levels and SARS-CoV-2 viral dynamics : implications to define correlates of protection against infection

**DOI:** 10.1101/2023.03.05.23286816

**Authors:** Guillaume Lingas, Delphine Planas, Hélène Péré, Darragh Duffy, Isabelle Staropoli, Françoise Porrot, Florence Guivel-Benhassine, Nicolas Chapuis, Camille Gobeaux, David Veyer, Constance Delaugerre, Jérôme Le Goff, Prunelle Getten, Jérôme Hadjadj, Adèle Bellino, Béatrice Parfait, Jean-Marc Treluyer, Olivier Schwartz, Jérémie Guedj, Solen Kernéis, Benjamin Terrier

**Author notes:** These authors contributed equally to this work as co-senior authors (Solen Kernéis, Benjamin Terrier).

## Abstract

**Background:** While anti-SARS-CoV-2 antibody kinetics have been well described in large populations of vaccinated individuals, we still poorly understand how they evolve during a natural infection and how this impacts viral clearance.

**Methods:** For that purpose, we analyzed the kinetics of both viral load and neutralizing antibody levels in a prospective cohort of individuals during acute infection by Alpha variant.

**Results:** Using a mathematical model, we show that the progressive increase in neutralizing antibodies leads to a shortening of the half-life of both infected cells and infectious viral particles. We estimated that the neutralizing activity reached 90% of its maximal level within 8 days after symptoms onset and could reduce the half-life of both infected cells and infectious virus by a 6-fold factor, thus playing a key role to achieve rapid viral clearance. Using this model, we conducted a simulation study to predict in a more general context the protection conferred by the existence of pre-existing neutralization, due to either vaccination or prior infection. We predicted that a neutralizing activity, as measured by ED_50_ >10^3^, could reduce by 50% the risk of having viral load detectable by standard PCR assays and by 99% the risk of having viral load above the threshold of cultivable virus.

**Conclusions:** This threshold value for the neutralizing activity could be used to identify individuals with poor protection against disease acquisition.

## Introduction

The analysis of viral and immunological kinetics during severe acute respiratory coronavirus 2 (SARS-CoV-2) infection has provided important insights on some patterns of the virus, both at the individual (within-host) and population (between-host) levels. For instance, we and others have found that SARS-CoV-2 peak viral load was close or even coincided with the onset of symptoms, suggesting that identifying individuals before symptoms onset was key to efficiently reduce transmission^1^. Likewise, we and others identified that dynamics of viral load after the peak was associated with the risk of severe disease, and we used these predictions to quantify the clinical efficacy of antiviral strategies^1, 2^. In addition, mathematical models of antibody kinetics after vaccination also played a key role to identify correlates of protection against severe infection^3^.

A question that has remained largely unsolved is the impact of antibody kinetics on viral clearance, and how the induction of antibodies modulates the time to viral clearance. Because the virus constantly mutates, it has been shown in large observational studies that the measurement of total anti-Spike (S) IgG antibodies was important^4–6^, but that their neutralization capacity was also critical^7^. Neutralization titers (ED50; half-maximal effective dilution), provides a much more accurate description of the quantitative and qualitative level of protection of patients’ sera, against the different Variants of Concerns (VoC) that emerged since 2021. This approach has been extensively used to analyze the magnitude and the duration of the protection conferred by mRNA vaccines^8^, and has played an important role to support booster dose strategies, or alert on the low level of protection of mRNA vaccine against disease acquisition in the Omicron variant era^9, 10^.

However, the combined kinetic analysis of both viral dynamic and neutralizing activity has never been studied in detail in the context of an acute infection. Here, we relied on the AMBUCOV cohort, a cohort of ambulatory individuals that took place in 2021 during the Alpha variant wave in France, prior to the mass vaccination campaign. Individuals were included early after symptoms onset, and both virological and immunological parameters were followed prospectively. We provided a detailed picture of the kinetics of antibody neutralization capacity against the Alpha variant, but also against the VoC that emerged subsequently, including Delta and Omicron (BA.1) variants. Following previous studies in hospitalized patients^11^, we used mathematical modeling to characterize the impact of the evolution of the neutralization activity on viral kinetics after a natural infection. Then we used this model to predict how the presence of a pre-existing neutralization activity (such as conferred by natural infection or vaccination) may reduce viral replication. We put these results in perspective to discuss the efficacy of vaccines and more broadly the use of neutralizing titers as a correlate of protection against disease acquisition.

## Patients and methods

### Study design

The AMBUCOV study (APHP201285, N° IDRCB /EUDRACT: 2020-A03102-37, ClinicalTrial.gov: NCT04703114) is a non-interventional longitudinal study that included 63 individuals between 05 February 2021 and 20 May 2021 in Cochin Hospital (Paris, France). The AMBUCOV study was an ancillary study of the cross-sectional SALICOV study (NCT04578509), that aimed to compare diagnostic accuracy of two alternate diagnosis strategies (nasopharyngeal antigen test and saliva nucleic acid amplification testing) to the current reference standard (nasopharyngeal nucleic acid amplification testing) for detection of SARS-CoV-2 in community testing centers^12^. The SALICOV study was conducted in the network of community screening centers of the Assistance Publique-Hôpitaux de Paris (APHP), France. Briefly, all individuals with symptoms (i.e., temperature > 37.8 °C or chills, cough, rhinorrhea, muscle pain, loss of smell or taste, unusual persistent headaches, or severe asthenia) were invited to be tested for SARS-CoV-2 in two community screening centers located in Paris . Testing was also available to all asymptomatic individuals wishing to be tested (i.e., contact of infected cases, before or after travel, after participation to a gathering event). Once their participation to SALICOV was completed, participants tested positive for SARS CoV-2 were contacted by phone by the principal investigator (BT) to explain the study protocol and offered to participate in the AMBUCOV study. Home visits were organized and written informed consent was obtained from all included participants (or their legal representatives if unable to consent).

Exclusion criteria included patients with criteria for hospitalization at the time of diagnosis, non-consent or inability to obtain consent, patients with dementia or not authorized, for psychiatric reasons or intellectual failure, to receive information on the protocol and to give informed consent, and patients under guardianship or curatorship.

### Study population and procedures

All adults included in the SALICOV study, with a positive nasopharyngeal PCR for SARS-CoV-2 within 48 hours, either with or without symptoms were included in the AMBUCOV study.

For each participant, four home visits were done by study nurses on day 0 (defined as the first study visit), day 3, day 8 and day 15. Blood samples were collected at each home visit, saliva on day 3, day 8 and day 15, nasal swab on day 8 and day 15 and stools on day 3 and day 15.

A follow-up study was performed at Cochin Hospital (Paris, France) on day 90 to collect outcome data and additional biological samples (blood, saliva and stools). Saliva samples were self-collected under supervision of the nurse or the principal investigator. Blood samples, saliva and stools samples were centralized, frozen in several aliquots at − 80°C within 24 hours and stored for analysis.

### Data collection

We collected data on sociodemographic, past medical history, presence of symptoms and concomitant medications using a standardized data collection form. When missing, date of symptom onset was imputed at the median observed in the population.

### Institutional Review Board (IRB) approval

The IRB C.P.P. Ile de France III approved the study protocol prior to data collection (approval number Am8849-2-3853-RM) and all subsequent amendments.

### Quantification of SARS-CoV2 RNA in saliva samples

Viral RNA was extracted from saliva samples using the Cellfree200 V7 DSP 200 protocol with the QIAsymphony® DSP virus/pathogen mini kit (QIAGEN, UK). Samples loaded onto the QIAsymphony® SP as instructed by the manufacturer, with a 200 μl sample input volume and 60 μl elution output volume of AVE buffer, unless stated (QIAGEN, UK). SARS-CoV-2 RT-ddPCR assays were performed using the One-Step RT-ddPCR Advanced Kit for 90 Probes (Bio-Rad Laboratories, Hercules, CA, USA) and the QX200 ddPCR platform (Biorad). A 2-plex RT-ddPCR assay was developed, which targets the Nucleocapside (N1) gene of the SARS-CoV-2 positive-strand RNA genome with specific FAM-probe and primers Cy5-labeled probe for the detection of a human housekeeping gene (RNAseP). RNAseP positivity was necessary to validate the RT-PCR assay prior to any further analysis. We considered 6 log_10_ copies/mL as a proxy for positive viral culture^11^.

### S-Flow Assay

The S-Flow assay is based on the recognition of the SARS-CoV-2 spike protein expressed on the surface of 293T cells. It was used to quantify SARS-CoV-2-specific IgG and IgA subtypes in sera as previously described^13, 14^. Briefly, 293T cells were obtained from ATCC (ATCC Cat# CRL-3216, RRID:CVCL_0063) and tested negative for mycoplasma. 293T cells stably expressing Spike (293T S) or control (293T Empty) were transferred into U-bottom 96-well plates (10^5^ cells/well). Cells were incubated at 4°C for 30 min with serum (1:300 dilution), saliva (1:5 dilution) or nasopharyngeal swabs (1:5 dilution) in PBS containing 0.5% BSA and 2 mM EDTA. Then, cells were washed with PBS, and stained at 4°C for 30 min using anti-IgG AlexaFluor647 (Jackson ImmunoResearch cat# 109-605-170) and Anti-IgA AlexaFluor488 (Jackson ImmunoResearch cat# 109-545-011). Then, cells were washed with PBS and fixed 10 min with 4% PFA. Data were acquired on an Attune Nxt instrument (Life Technologies). Results were analyzed with FlowJo 10.7.1 (Becton Dickinson). The specific binding was calculated as follow: 100 x (% binding 293T Spike - % binding 293T Empty)/ (100 - % binding 293T Empty). For sera, the assay was standardized with WHO international reference sera (20/136 and 20/130) and cross-validated with two commercially available ELISA (Abbott and Beckmann) using a Passing-Bablok linear regression model to allow calculation of BAU/mL^15^. SARS-CoV-2 neutralization was assessed using the S-fuse assay, as previously described^16^.

### S-Fuse neutralization assay

U2OS-ACE2 GFP1-10 or GFP 11 cells, also termed S-Fuse cells, become GFP+ when they are productively infected by SARS-CoV-2^17^. Cells tested negative for mycoplasma. Cells were mixed (ratio 1:1) and plated at 8 × 10^3^ per well in a μClear 96-well plate (Greiner Bio-One). The indicated SARS-CoV-2 strains were incubated with serially diluted sera for 15 min at room temperature and added to S-Fuse cells. Sera were heat-inactivated for 30 min at 56 °C before use. 18 h later, cells were fixed with 2% PFA, washed and stained with Hoechst (dilution of 1:1,000, Invitrogen). Images were acquired using an Opera Phenix high-content confocal microscope (PerkinElmer). The GFP area and the number of nuclei were quantified using the Harmony software (PerkinElmer). The percentage of neutralization was calculated using the number of syncytia as value with the following formula: 100 × (1 – (value with serum – value in ‘non-infected’)/(value in ‘no serum’ – value in ‘non-infected’)). Neutralizing activity of each serum was expressed as the half maximal effective dilution (ED50).

#### Viral strain

B.1.1.7 (Alpha variant) was isolated from an individual in Tours (France) who had returned from the UK (PMID: 33772244). B.1.617.2 (Delta variant) was isolated from a nasopharyngeal swab of a hospitalized patient who had returned from India. The swab was provided and sequenced by the Laboratoire de Virologie of the Hôpital Européen Georges Pompidou (Assistance Publique des Hôpitaux de Paris) (PMID: 34237773). The Omicron BA.1 strain was supplied and sequenced by the NRC UZ/KU Leuven (Leuven, Belgium) (PMID: 35016199).

All individuals provided informed consent for the use of the biological materials. The variant strains were isolated from nasal swabs on Vero cells and amplified by one or two passages on Vero cells.

Titration of viral stocks was performed on Vero E6, with a limiting dilution technique allowing a calculation of TCID50, or on S-Fuse cells. Viruses were sequenced directly on nasal swabs, and after one or two passages on Vero cells. Sequences were deposited in the GISAID database immediately after their generation, with the following IDs: Alpha: EPI_ISL_735391; Delta: ID: EPI_ISL_2029113; Omicron BA.1.

### Model for antibody and ED_50_ kinetics

We modeled the evolution of IgG levels using a sigmoid Gompertz function to reflect the progressive increase in IgG from 0 (before infection) to a plateau, noted IgG_max_:

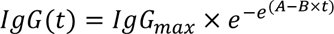

We next relate IgG to the evolution of the neutralizing activity (*ED_50_*) against different strains, namely Alpha (α), Delta (δ) and Omicron (BA.1, ο) using the following relationship:

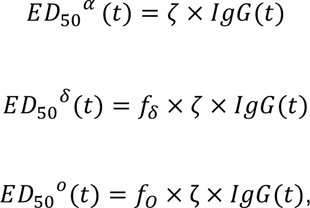

such that represents the scaling factor between IgG and 𝐸𝐷_50_^𝛼^, while f (resp f) represent the fold change between the neutralization capacity against alpha and delta variant (resp. omicron). Of note, in this model, the time to reach 90% of the maximal protection is the same for all variants and is equal to (A - log(-log(0.9))/B).

### Model for viral dynamics in saliva

We used a target-cell limited model with an eclipse phase as described before^11^ **(Supplementary Figure 1)** to characterize viral dynamics in saliva from infection (t=0) to clearance. In brief, the model includes three types of cell populations: target cells (T), infected cells in an eclipse phase (I_1_) and productively infected cells (I_2_). The model assumes that target cells are infected at a constant infection rate 𝛽 (mL.virion^-1^.d^-1^). Once infected, cells enter an eclipse phase and become productively infected after a mean time 1/k (day). We assume that productively infected cells have a constant loss rate, noted 𝛿 (d^-1)^. Infected cells produce p viral particles per day (virus.d^-1^), but only a fraction of them, 𝜇, is infectious, and the virus particles can either be infectious (V_I_) or non-infectious (V_NI_). We assumed that viral load, as measured by RNA copies (V), is the sum of infectious and non-infectious viral particles, both cleared at the same rate, c. The model can be written as :

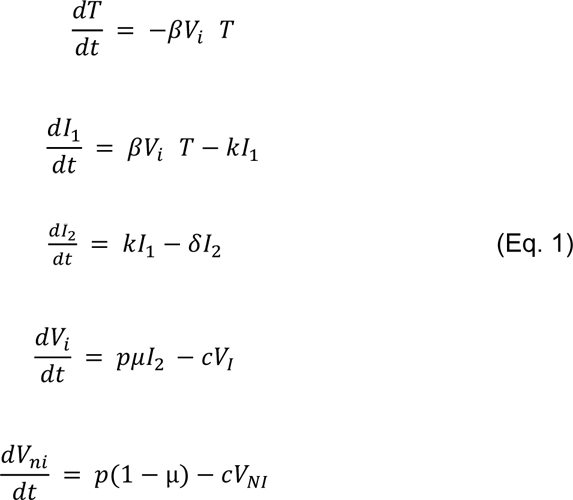

The basic reproductive number R_0_, defined by the number of secondary infected cells resulting from one infected cell in a population of fully susceptible cells, T_0_, is defined by :

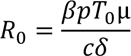

### Combined immunovirological model

Finally, we aimed to characterize the impact of the neutralizing antibody level on viral load. For that purpose, we tested several models, assuming no effect of neutralization antibody levels (model M0, Eq. 1), or that the effect of neutralization could alternatively i) increase infected cell clearance (model M1), ii) increase the loss of both infectious and non-infectious virus (model M2), iii) or both (model M3) (**Supplementary Figure 1**).

Model M0:

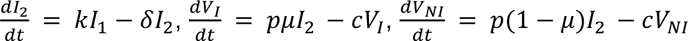

Model M1:

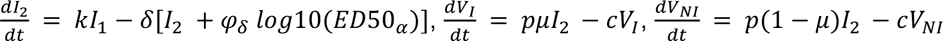

Model M2

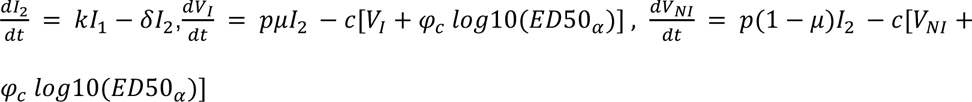

Model M3:

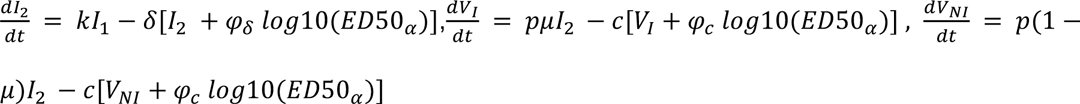

### Assumptions on parameter values

We fixed c to 10 d^−1^, k to 4 d^−1^ and μ to 10^−4^ as previously published^11^. As only the product p×T_0_ is identifiable, we also fixed the density of susceptible epithelial cells to the same value found in the upper respiratory tract, i.e., T_0_= 1.33×10^5^ cells.mL^-1^. Further we assumed that the duration of the incubation period was log-normally distributed, with a median value of 5 days a standard deviation of 0.125, such that 90% of individuals have an incubation period between 3 and 7 days^11^. Thus, only 3 viral parameters were estimated, namely p, δ and R_0_, along with their interindividual variabilities. Given the lack of data on the viral upslope, we also fixed the standard deviation of the random effect associated to R_0_, denoted ω_R0_ to 0.5, as done previously^18^.

### Inference & model selection

Models M0, M1, M2, and M1+M2 (i.e., a dual effects on both infected cells and virus clearance) were fitted to all data available, namely viral load, IgG and 𝐸𝐷_50_ against all strains, assuming an additive error on the log-quantities. Parameters were estimated using non-linear mixed effect models and SAEM algorithm, using the same statistical methodology as previously described^11, 18, 19^. Only the results obtained with the best model are presented.

### Impact of a pre-existing neutralization capacity on viral dynamics

Next, we used the best model to anticipate the viral dynamics that could be observed in non-naive individuals, i.e., in individuals having a pre-existing neutralization due either natural infection or vaccination. For that purpose, we assumed that one virus was present at t=0 (infection time), and we assumed different levels of neutralizing capacity ranging from ED_50_=0 to ED_50_=10^5^. For each value of ED_50_ we generated a large population of 5,000 virological profiles using the final immuno-virological model, and we calculated different viral metrics. Of note, we made the conservative assumption here that the neutralizing capacity remained constant during the infection, i.e., we did not consider any increase over time due to stimulated immune response. As a sensitivity analysis, we also calculated the protection obtained with the alternative models.

#### Materials availability statement

All codes, datasets for the modelling analysis and datasets for the figures, supplementary figures, tables and supplementary tables are available at https://datadryad.org/stash/share/oNDQyb2rNuSRXr8vjIZSHBtIIC6zp4h504gunxxjmbo.

## Results

### Baseline characteristics

A total of 57 patients were included between February and September 2021 (**Table 1**). Patients were mostly male (*N* = 40, 63%), with a median age of 44 years (IQR: 35-57) and all were infected with the Alpha variant. Fifty-five participants developed symptoms, and 2 remained asymptomatic throughout the study. Patients had very few comorbidities, and hypertension (5%), chronic cardiac disease (5%), obesity (3%), and chronic kidney disease (2%) were the most common comorbidities. One patient was fully vaccinated (2 doses) and 7 patients had received one dose of vaccination at the time of infection. The median time between symptoms onset and inclusion in the AMBUCOV study was 4 (IQR: 3-6) days and the median saliva viral load at inclusion was 6.27 (IQR: 5.61-6.93) log_10_ copies/mL.

**Table 1.**
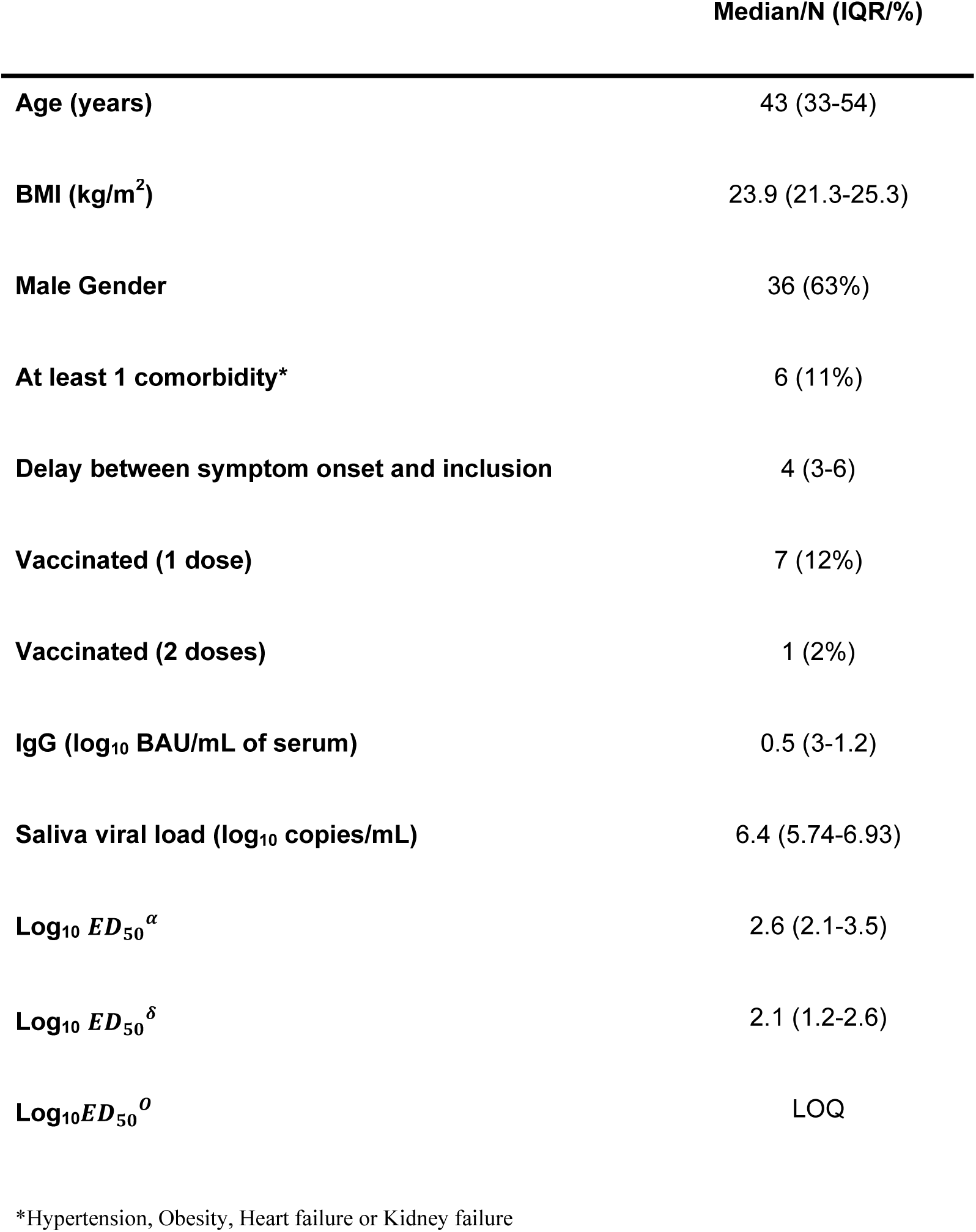
Clinical and biological characteristics at inclusion in the AMBUCOV study.

### Immuno-virological modeling

All data used for the modeling exercise, namely viral load (in saliva), anti-S IgG and neutralizing titers (in plasma or serum) are shown in **Figure 1** and **Supplementary Figure 2**.

**Figure 1.**
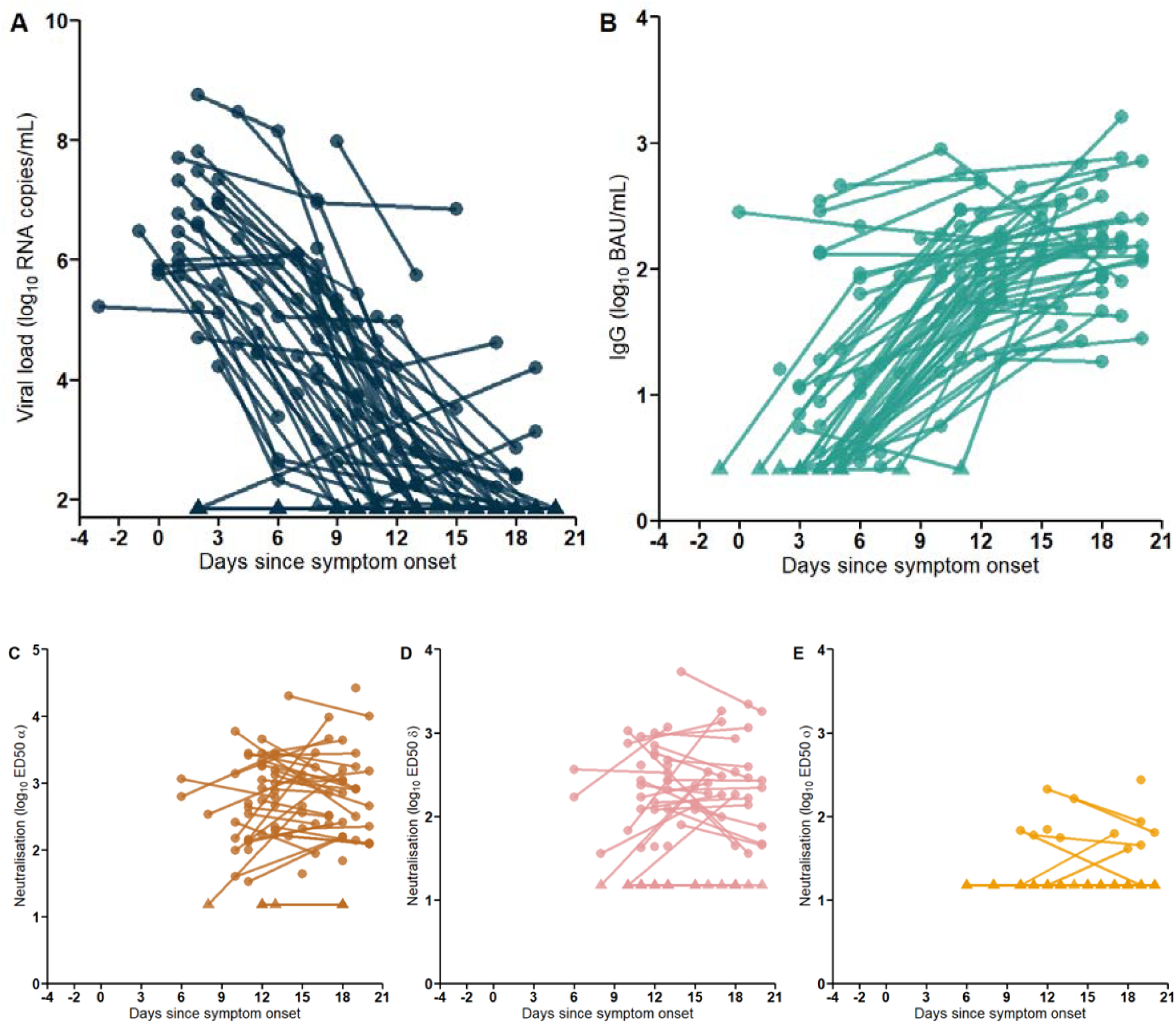
Virological and immunological data analyzed in the AMBUCOV cohort. A. Saliva viral load. B. Serum concentration of IgG (BAU/mL). C-E. Neutralization activity of IgG against strains C. Alpha D. Delta. E. Omicron (BA.1). All data expressed in time since symptom onset. Triangles represent data below LOQ.

The model best describing our data assumed that neutralizing antibodies acted on both infected cell and infectious virus clearance (M1+M2), and the model could well fit all data (**Figure 2**). Model parameters were in line with what we found in other studies^1, 18,,19^, with a within-host R_0_ equal to 22.6, a viral production rate of 4 x 10^3^ viruses/cell/day, and a loss rate of infected cells in absence of antibodies equal to 𝛿 = 0.26 𝑑^−1^ (**Table 2**). The peak viral load occurred at symptom onset with a median level of 6.8 log_10_ RNA copies/mL.

**Figure 2.**
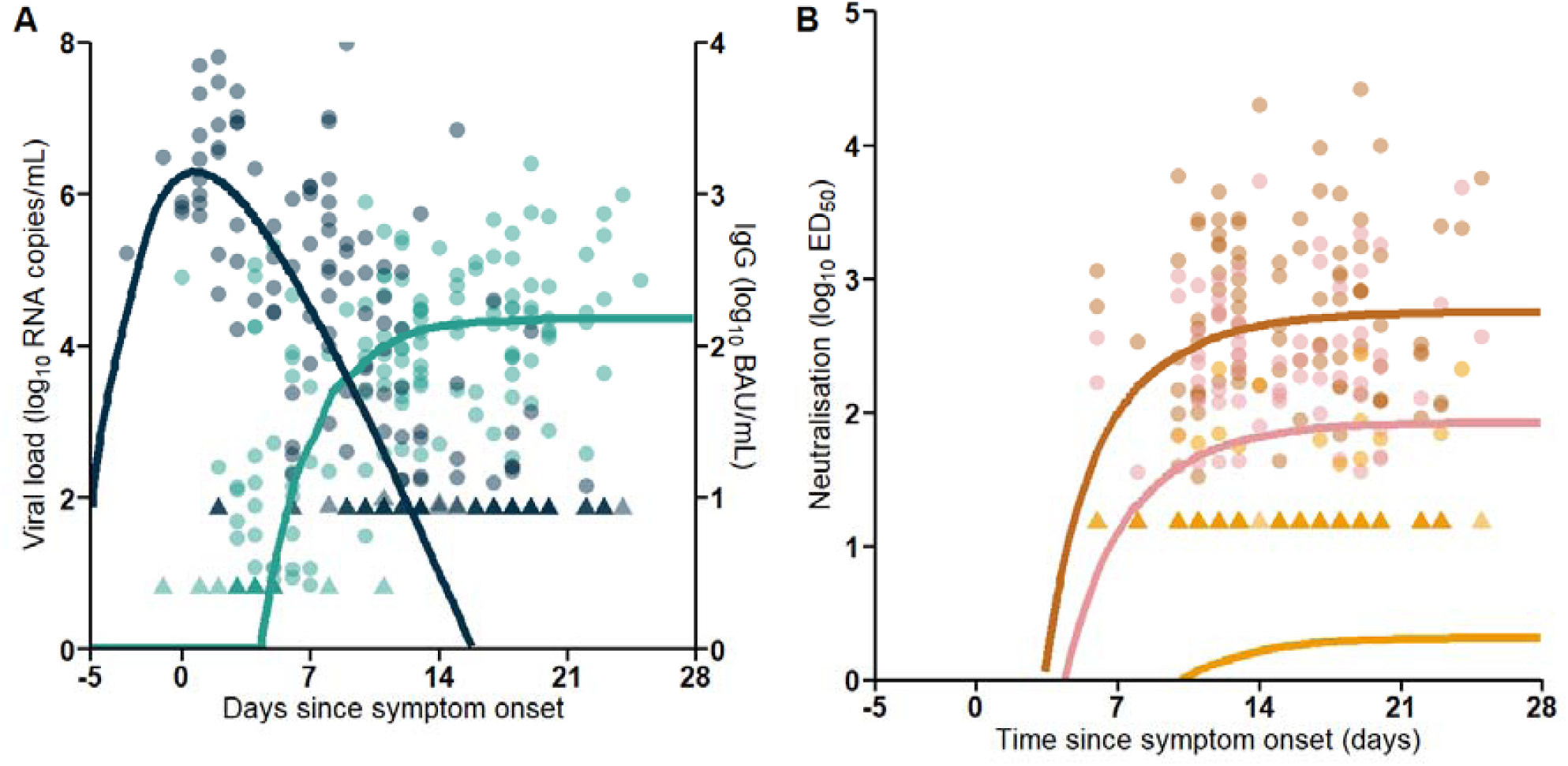
Median predictions of viral (A) and serological (B) kinetics. Circles are the observed data and lines represent the simulation-based median predictions of the model. Triangles represent data below LOQ. Darkblue: Viral load. Lightblue: IgG. Brown: ED_50_ɑ. Pink: ED_50_δ. Yellow: ED_50_ο

**Figure 3.**
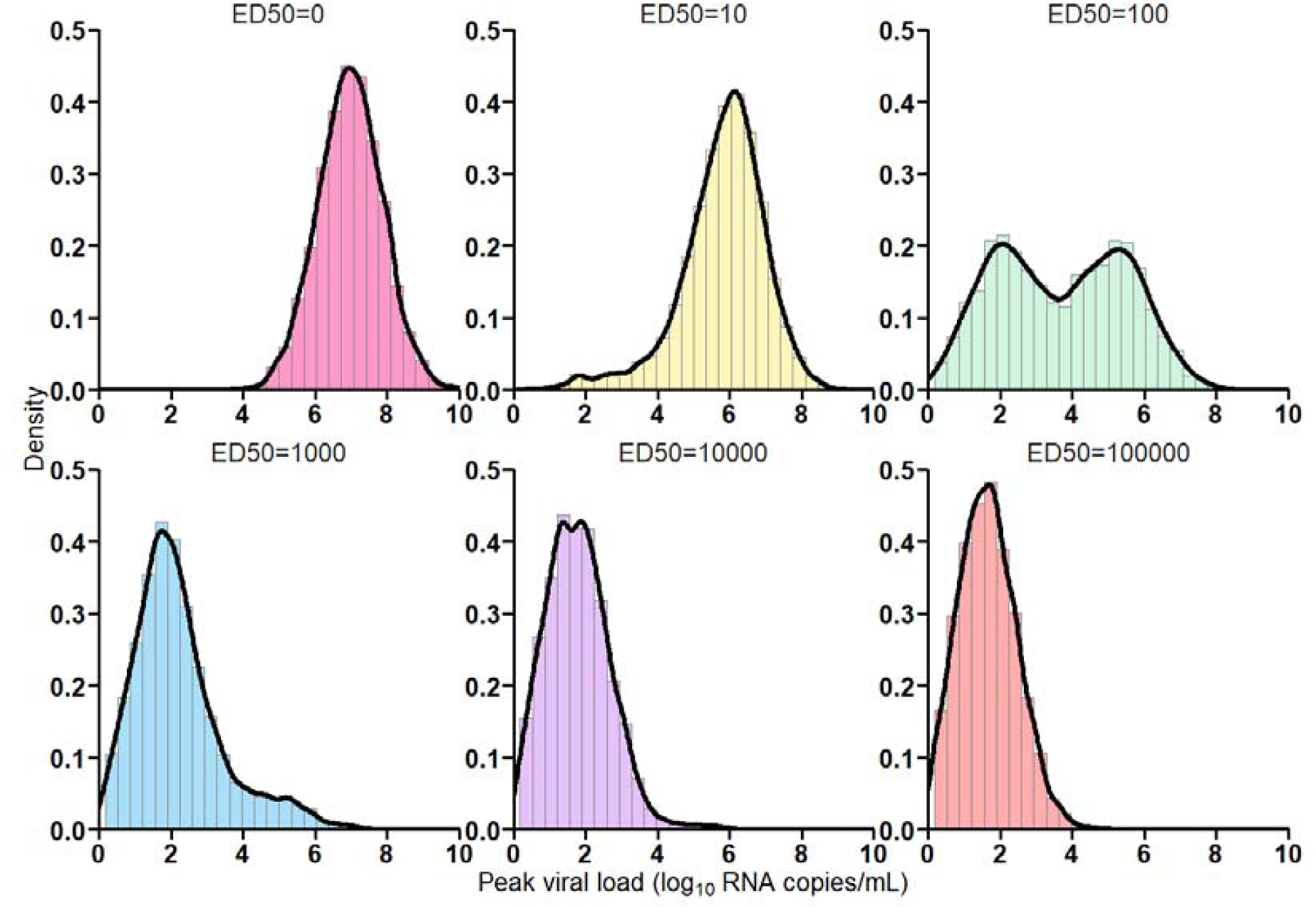
Prediction of peak viral load distribution depending on ED_50_ levels at initiation of infection. Values of ED_50_ : Pink: 0 ; Yellow: 10 ; Green: 100 ; Blue : 1000 ; Purple : 10000 ; Red : 100000

**Table 2.** Predicted impact of a pre-existing antibody neutralization on viral kinetics. The limit of detection and the threshold of infectivity were set to 1.84 and 6 log_10_ copies/mL, respectively.

| Antibody neutralization level at infection (ED<sub>50</sub>) | Median peak viral load (log<sub>10</sub> copies/mL) | Probability of peak viral load above the limit of detection | Probability of peak viral load above the threshold of infectivity |
| --- | --- | --- | --- |
| 0 | 7.2 | 100% | 86% |
| 10<sup>1</sup> | 6.0 | 99% | 48% |
| 10<sup>2</sup> | 4.1 | 80% | 12% |
| 10<sup>3</sup> | 1.9 | 55% | 1% |
| 10<sup>4</sup> | 1.7 | 44% | 0% |
| 10<sup>5</sup> | 1.6 | 38% | 0% |

The population average maximal level of anti-S IgG after acute infection, IgG_max_, was equal to 155 BAU/mL, corresponding to an antibody neutralization level against 𝛼 variant, ζ* IgG_max_, equal to 548 ED_50_. After infection, antibodies rapidly increased, and we predicted that 90% of this maximal antibody protection was achieved as early as day 10 post-symptom onset. This level of neutralization was achieved around day 6 after symptoms onset in patients vaccinated with one dose, and the only patient that had received 2 doses at the time of the infection reached this level only 4 days after symptoms onset, supporting that vaccination considerably reduced the time to achieve high level of neutralization activity. At antibody peak, we estimated that the half-lives of both infected cells and infectious virus were shortened by respectively 6- and 7-fold (corresponding to loss rates for 𝛿 and c equals to 1.56 d^-1^ and 77 d^-1^, respectively). Because antibody levels reached their maximal value after peak viral load, we did not find a significant association between the cumulated levels of neutralizing antibody levels and viral load (**Supplementary Figure 3**). As all individuals were infected with Alpha variant, the population average maximal level of neutralization against Delta and BA.1 variants were much lower and were diminished by respectively 6.7- and 277.7-folds (ζ*IgG_max_*f_Delta/Omicron_), leading to median ED_50_ of 82 and 2, respectively, after infection, reached around 19 days after symptom onset.

To address the impact of the temporal effect of antibody levels on viral clearance, we simulated 5000 *in silico* virological profiles using the estimated parameter distributions and considering that antibody could have either the two mechanisms of action (as found in our model), only one of them or none of them (thus fixing alternatively 𝜑_ð_ and/or 𝜑_𝑐_ to 0 in the model). When considering the full model, the predicted median time to clearance after symptoms onset was equal to 12 days, as compared to >50 in a model in which antibodies had no effect (𝜑_ð_ = 𝜑_𝑐_ = 0). We observed that the effectiveness of IgG was predominantly driven by its action on the loss rate of infected cells, with a median time to viral clearance equal to 14 days when only the effects on infected cell was assumed (𝜑_𝑐_ = 0) as compared to >50 days when only the effects on infected viral particles was assumed (𝜑_ð_ = 0) (**Supplementary Figure 4**). Consistent with this prediction, the post-hoc analysis showed that the early appearance of detectable neutralizing antibodies was associated with lower viral levels at day 4 post-symptom onset, which corresponds to the median time of inclusion in the study (r=0.47, (P<10^-3^, **Supplementary Figure 3**).

### Impact of a pre-existing neutralization capacity on viral dynamics

Next, we used the model to anticipate the viral dynamics that could be observed in non-naive individuals after an encounter with the virus, i.e., in individuals having a pre-existing neutralization due either natural infection or vaccination. For that purpose, we assumed that infection is initiated at t=0 with only one infectious particle, and we assumed different levels of neutralizing capacity ranging from ED_50_=1 to ED_50_=10^5^ (see methods). This corresponds to a within-host R_0_ ranging from 22.5 (i.e., the value estimated in our population before antibody secretion) to about 0.5. Using the model parameters, we simulated viral dynamics of 5,000 individuals with each potential level of ED_50_ and we computed the following metrics: peak viral load, probability of having detectable viral load at peak, probability of having viral load > 6 log_10_ copies/mL. The simulations showed that ED_50_>1000 would be sufficient to maintain 45% of individuals with viral load below the limit of detection at all times, and only 1% being at risk of transmitting the infection (i.e.peak viral load above > 6 log_10_ copies/mL).

## Discussion

In this work, we combined the kinetic analysis of saliva viral load and immune response during an acute Covid-19, from infection to viral clearance in ambulatory patients with non severe disease. We show that neutralizing antibodies on infected cells and, to some extent, on circulating viral particles, played a key role to achieve viral clearance. The neutralizing activity was largely variant-dependent, and ED_50_ was estimated to be equal to 548 against Alpha variant but decreased by 7- and 300-fold against Delta and Omicron BA.1 variants, respectively. We next performed simulations to predict the level of protection against infection conferred by various pre-existing levels of antibody neutralization, and predicted that a level of ED_50_ >10^3^ was sufficient to prevent 50% of infections from being detectable and 99% from being above the threshold of viral culture, used as a proxy of infectiousness.

The AMBUCOV study population included patients with mild COVID-19 during the Alpha variant wave in France, prior to the mass vaccination campaign, to describe the natural course of viral load and immune response in immunocompetent patients without any strong comorbidities, as illustrated by patients characteristics. The decision was directly related to our main objective, i.e. to analyze the relationship between the virus and the immune response.

We have modeled the kinetics of both saliva viral load and immune response, mainly the humoral response, during an acute COVID-19. We showed that the increase in neutralizing antibodies leads to a shortening of the half-life of both infected cells and infectious viral particles. We estimated that the neutralizing activity reached 90% of its maximal level within 8 days after symptoms onset and could reduce the half-life of both infected cells and infectious virus by a 6-fold factor, thus playing a key role to achieve rapid viral clearance. To establish a correlate of protection against SARS-CoV-2 infection, we predicted that a neutralizing activity defined by ED_50_ >10^3^ could reduce by 50% the risk of having viral load detectable in saliva by ultrasensitive ddPCR assays and by 99% the risk of having viral load above the threshold of cultivable virus. Overall, this value of neutralizing activity could be used to identify individuals with poor protection against disease acquisition.

Based on the data from a previous study^8^, we compared the level of neutralizing antibodies in individuals hospitalized in a nursing home prior to an outbreak, and compared the levels between individuals that experienced a breakthrough infection and those that remained uninfected. While all individuals in this study were vaccinated, the median ED_50_ before infection was 1429 in individuals that subsequently experienced a breakthrough infection (with Omicron BA.1 variant) as compared to 2528 in those who did not. Similarly, in two studies, levels of neutralizing antibodies were lower 1770 against Alpha and BA.5 variants just before the respective breakthrough infection^20, 21^. These data during the Omicron wave support our findings and the threshold of neutralizing activity ED_50_ >10^3^ as a potential correlate of protection against SARS-CoV-2 infection regardless of the variants.

Finally, our study has some limitations. Viral loads and IgG were not quantified from same site, the first being obtained in saliva and the second in serum. Unfortunately the neutralizing assay was not adapted to measure the neutralizing activity in saliva.Further, as frequently observed in acute infection^1, 18^, very few data could be measured before peak viral load, which may cause a bias in the estimation of R_0_ and, accordingly, their effects on loss rates parameters. Finally, our identifiability analysis showed that the effect neutralization of circulating viral particles 𝜑_𝑐_ was poorly identifiable (RSEE = 76%) and that the effect of neutralization on infected cells clearance rate was barely identifiable as well (RSEE = 49%). All identifiability metrics are available in Supplementary Table 4. As a consequence, we conducted the same 5000 individual simulations as previously, this time using the estimates of model M1, where only an effect on the clearance rate of infected cells was considered. In these simulations, we observed that above 10^3^ ED50, the resulting protection conferred did not improve (Supplementary table 5), with peak viral loads and proportion of detectable individuals at peak viral load being >98% at all concentrations.

In conclusion, our data show that ED_50_ >10^3^ could be a clinically relevant threshold value for the neutralizing activity to identify individuals with poor protection against disease acquisition. The evaluation of this threshold on larger cohorts is now warranted to evaluate whether it could be used to define a correlate of protection against disease acquisition.

## Data Availability

All codes, datasets for the modelling analysis and datasets for the figures, supplementary figures, tables and supplementary tables are available at .

---

**Supplementary figure 1.**
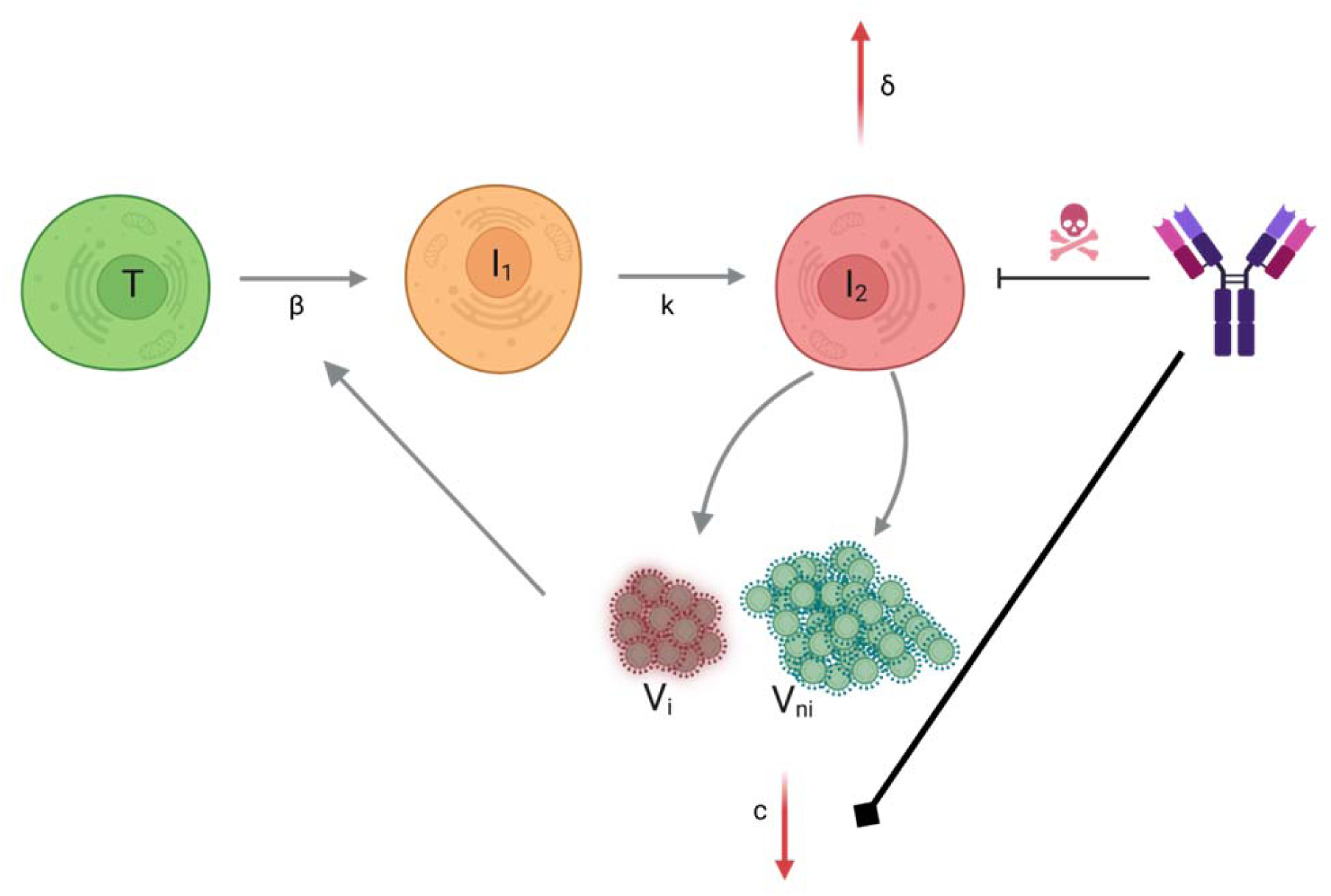
Viro-immunological model.

**Supplementary figure 2.**
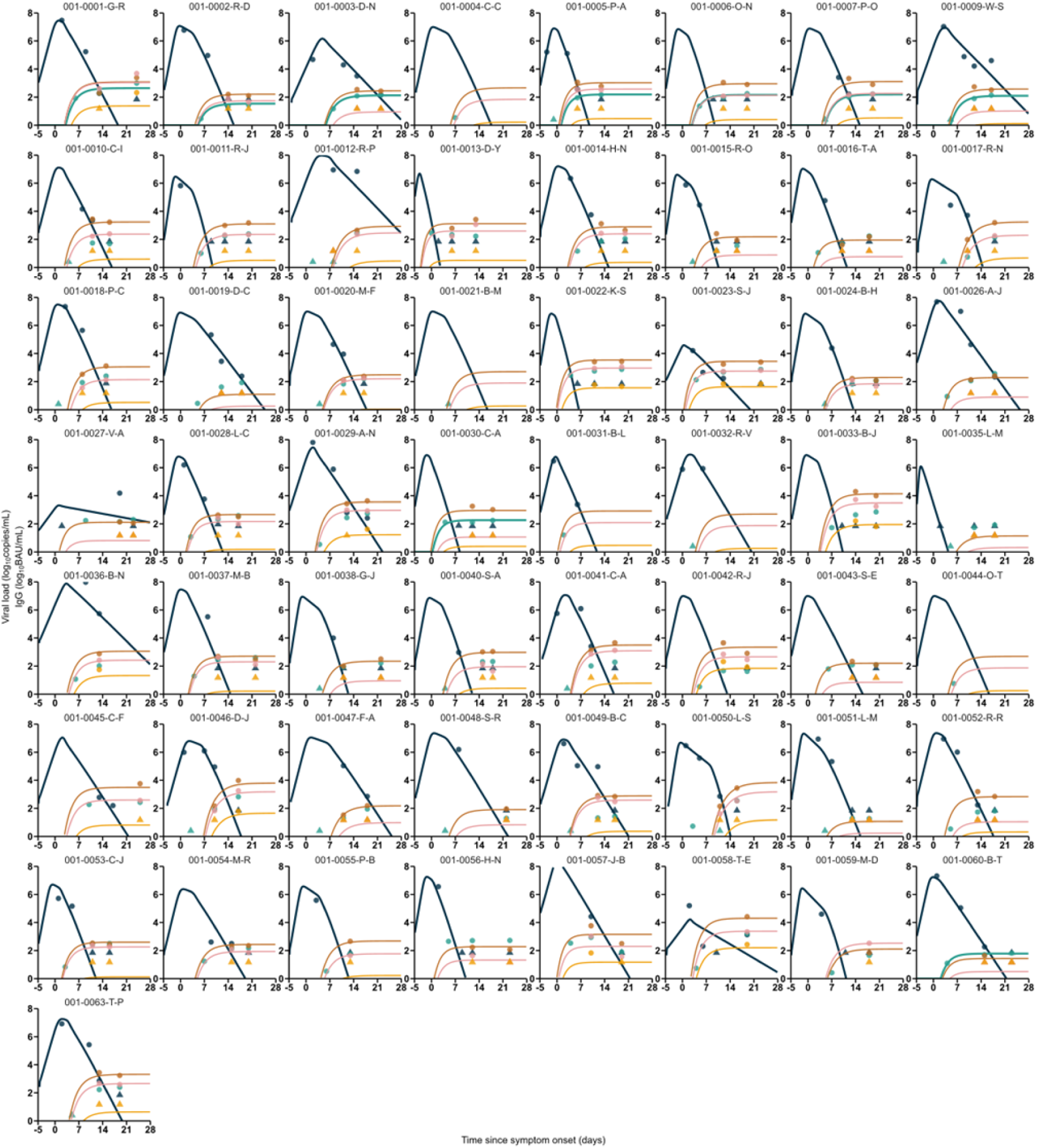
Individual fits of the whole population studied.

**Supplementary figure 3.**
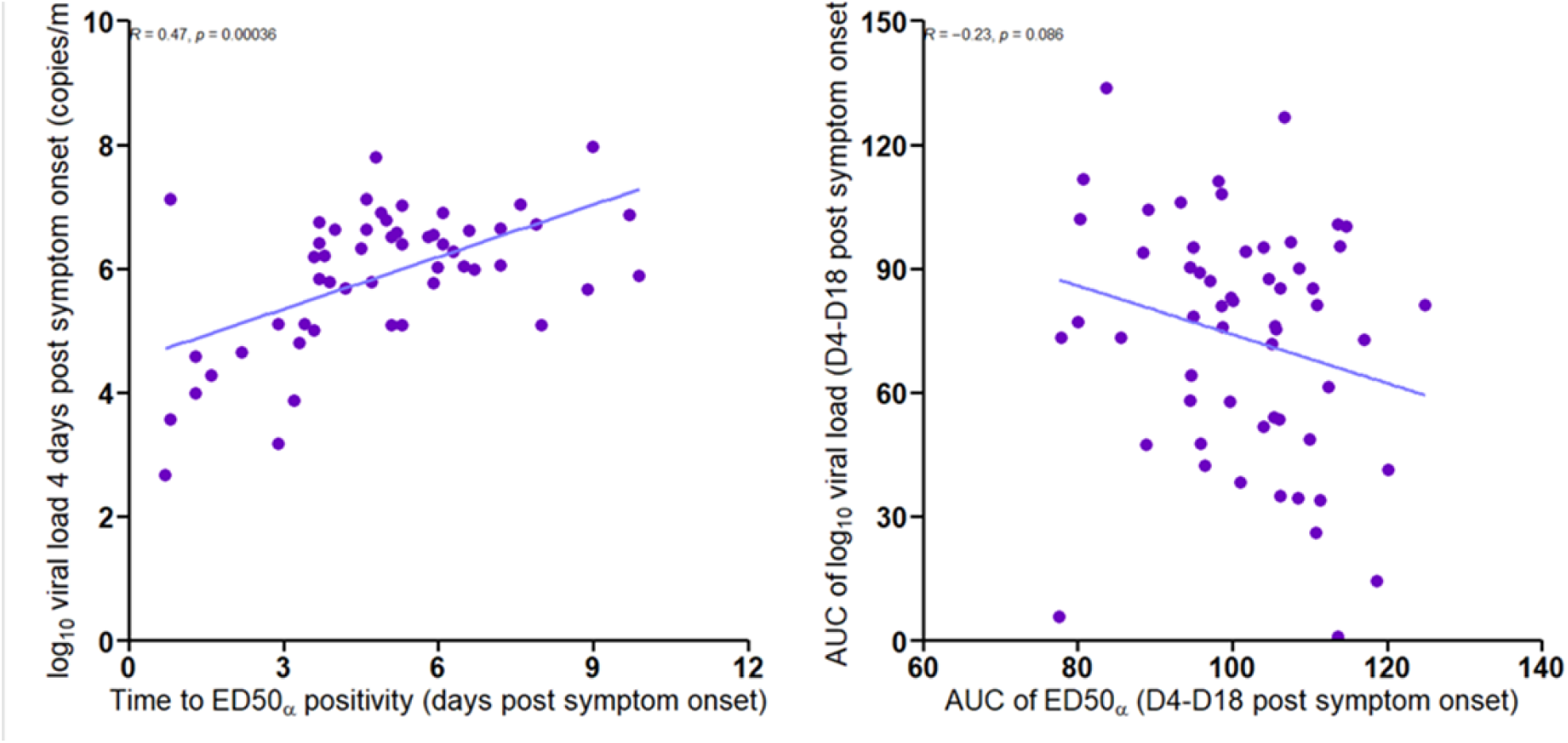
Correlations between predicted viral load 4 days post symptom onset and time to ED_50_ positivity (left) and AUCs of viral load and ED_50_ between 4 and 18 days after symptom onset.

**Supplementary figure 4.**
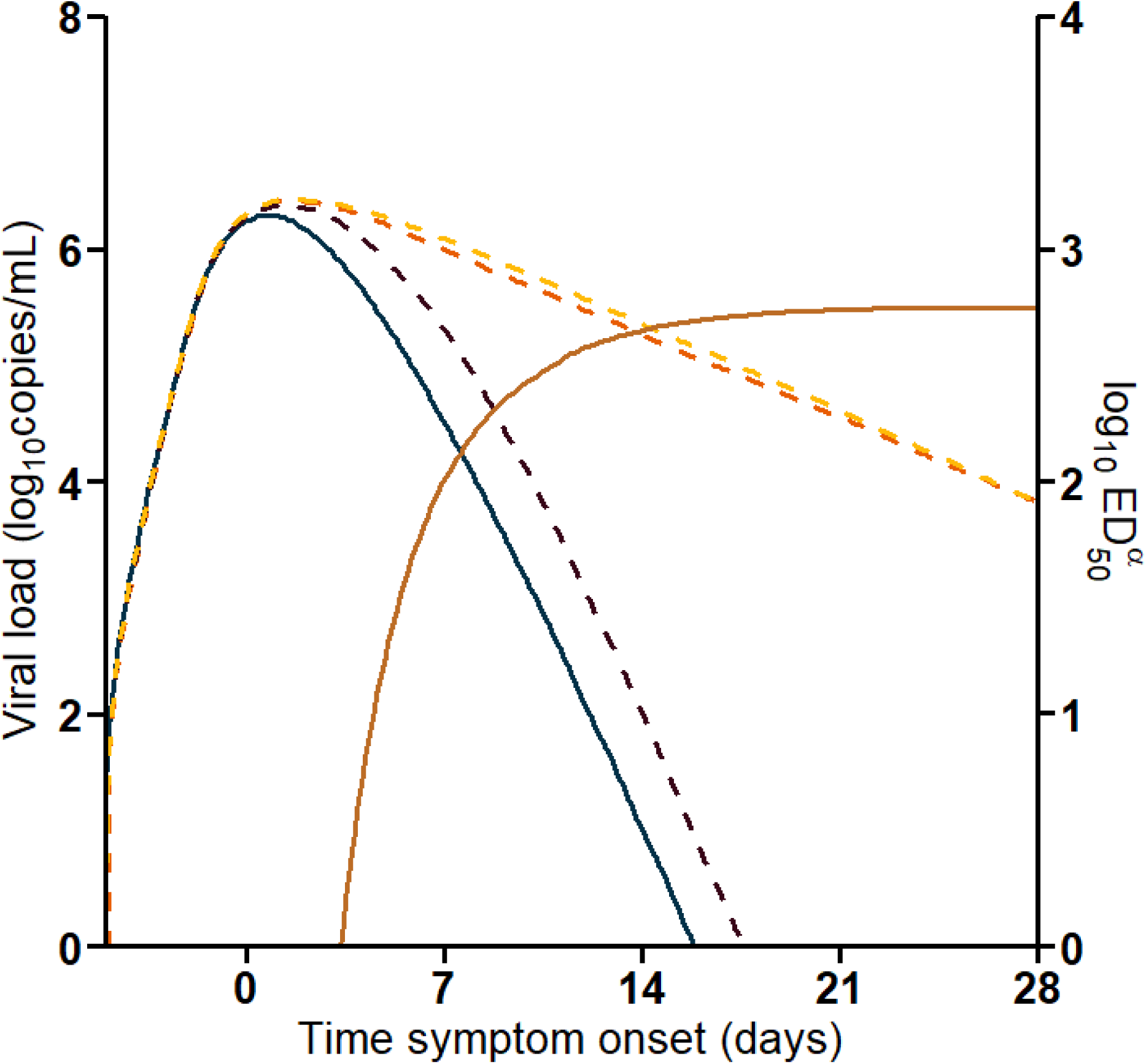
Sensitivity analysis of each mode of action of neutralization. Green: median predicted viral load of model M1+M2. Yellow: Clearance rate of viral particles not increased by neutralization. Red: Clearance rate of infected cells not increased by neutralization.

**Supplementary Figure 5.**
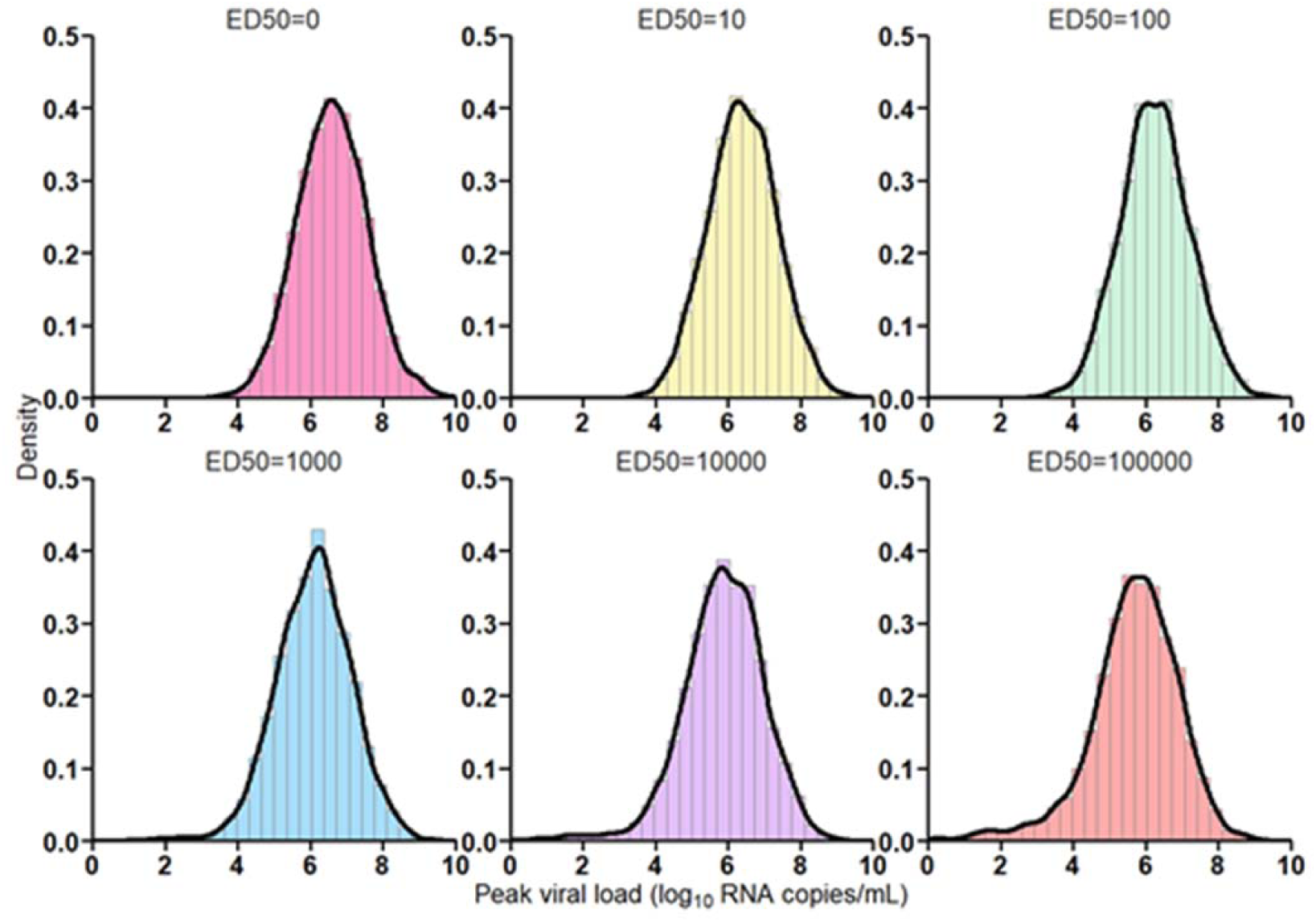
Prediction of peak viral load distribution depending on ED_50_ levels at initiation of infection using parameter estimates of model M1.

**Supplementary table 1.** Parameter estimates of model M0.

|  | Parameter estimate (RSE, %) |  |
| --- | --- | --- |
|  | Fixed effect | Random effect SD |
| **R0** | 19.1 (58.3) | 0.5 |
| **k (d<sup>-1</sup>)** | 4 | - |
| **p (virus/cell/d)** | 17704 (68.8) | 1.76 (46.7) |
| **c (d<sup>-1</sup>)** | 10 | - |
| **μ** | 0.0001 | - |
| **δ (d<sup>-1</sup>)** | 0.88 (7.32) | 0.28 (24.6) |
| **IgG<sub>max</sub>** | 161 (15) | 0.94 (12.0) |
| **A** | 4.63 (11.4) | 0.15 (50.9) |
| **B** | 0.36 (13.8) | 0.19 (46.7) |
| **ζ** | 3.58 (21) | 1.36 (12.3) |
| **f<sub>delta</sub>** | 0.15 (22.5) | 1.26 (17.2) |
| **f<sub>O</sub>** | 0.0037 (50.6) | 1.11 (42.8) |
| **φ<sub>δ</sub>** | 0 | - |
| **φ<sub>c</sub>** | 0 | - |
| **BIC** | 1173.09 |  |

**Supplementary table 2.** Parameter estimates of model M1.

| Parameter estimate (RSE, %) |  |  |
| --- | --- | --- |
|  | Fixed effect | Random effect SD |
| **R0** | 17.7 (58.3) | 0.5 |
| **k (d<sup>-1</sup>)** | 4 | - |
| **p (virus/cell/d)** | 1803 (68.2) | 2.23 (33.5) |
| **c (d<sup>-1</sup>)** | 10 | - |
| **μ** | 0.0001 | - |
| **δ (d<sup>-1</sup>)** | 0.35 (21.6) | 0.45 (28.9) |
| **IgG<sub>max</sub>** | 153.07 (14.7) | 12.5 (12) |
| **A** | 5.1 (8.58) | 0.19 (69.3) |
| **B** | 0.41 (9.86) | 0.12 (198) |
| **ζ** | 3.58 (20.1) | 1.35 (12.2) |
| **f<sub>delta</sub>** | 0.15 (23.2) | 1.32 (18.2) |
| **f<sub>O</sub>** | 0.0031 (109) | 1.27 (88.2) |
| **Φ<sub>δ</sub>** | 1.14 (10.8) | - |
| **Φ<sub>c</sub>** | 0 | - |
| **BIC** | 1158.54 |  |

**Supplementary table 3.** Parameter estimates of model M2.

| Parameter estimate (RSE, %) |  |  |
| --- | --- | --- |
|  | Fixed effect | Random effect SD |
| **R0** | 17.9 (100) | 0.5 |
| **k (d<sup>-1</sup>)** | 4 | - |
| **p (virus/cell/d)** | 15609 (72.1) | 2.03 (27.2) |
| **c (d<sup>-1</sup>)** | 10 | - |
| **μ** | 0.0001 | - |
| **δ (d<sup>-1</sup>)** | 0.88 (7.77) | 0.69 (16.8) |
| **IgG<sub>max</sub>** | 157.84 (14.7) | 0.92 (12) |
| **A** | 4.82 (9.56) | 0.13 (46) |
| **B** | 0.38 (11.6) | 0.2 (31.2) |
| **ζ** | 3.58 (21.2) | 1.37 (12.4) |
| **f<sub>delta</sub>** | 0.15 (23.0) | 1.3 (17.0) |
| **f<sub>o</sub>** | 0.0024 (85.5) | 1.5 (53.8) |
| **φ<sub>δ</sub>** | 0 | - |
| **φ<sub>c</sub>** | <10<sup>-5</sup> (>10<sup>3</sup>) | - |
| **BIC** | 1176.88 |  |

**Supplementary table 4.** Parameter estimates of model M1+M2.

|  | Parameter estimate (RSE, %) |  |
| --- | --- | --- |
|  | Fixed effect | Random effect SD |
| **R0** | 22.57 (33.5) | 0.5 |
| **k (d<sup>-1</sup>)** | 4 | - |
| **p (virus/cell/d)** | 4275.38 (63.2) | 2.03 (27.2) |
| **c (d<sup>-1</sup>)** | 10 | - |
| **μ** | 0.0001 | - |
| **δ (d<sup>-1</sup>)** | 0.26 (14.7) | 0.69 (16.8) |
| **IgG<sub>max</sub>** | 155 (14.5) | 0.94 (12) |
| **A** | 5.24 (11.3) | 0.12 (52) |
| **B** | 0.42 (14.2) | 0.21 (29.5) |
| **ζ** | 3.54 (20.9) | 1.34 (12.5) |
| **f<sub>delta</sub>** | 0.15 (22.7) | 1.27 (16.7) |
| **f<sub>o</sub>** | 0.0036 (81.6) | 1.22 (63.7) |
| **Φ<sub>δ</sub>** | 1.51 (3.90) | - |
| **Φ<sub>c</sub>** | 1.81 (2.47) | - |
| **BIC** | 1159.27 |  |

**Supplementary table 5.** Identifiability metrics of model M1+M2.

|  | Parameter estimate (RSE, %) | Empirical SE | Empirical RSE | RBias |
| --- | --- | --- | --- | --- |
| **R0** | 22.57 (33.5) | 14.8 | 65 | 25 |
| **p (virus/cell/d)** | 4275.38 (63.2) | 4971 | 116 | 41 |
| **$\delta$ (d<sup>-1</sup>)** | 0.26 (14.7) | 0.08 | 30 | -8.2 |
| **IgG<sub>max</sub>** | 155 (14.5) | 23 | 15 | -2.6 |
| **A** | 5.24 (11.3) | 0.6 | 11 | 2.3 |
| **B** | 0.42 (14.2) | 0.06 | 13.6 | 3.3 |
| **$\zeta$** | 3.54 (20.9) | 0.77 | 22 | 5.1 |
| **f<sub>delta</sub>** | 0.15 (22.7) | 0.04 | 26 | 5.1 |
| **f<sub>o</sub>** | 0.0036 (81.6) | 0.0018 | 50.0 | 8.4 |
| **$\phi_{\delta}$** | 1.81 (3.90) | 0.75 | 50 | 23.5 |
| **$\phi_c$** | 1.51 (2.47) | 1.37 | 76 | 23.1 |

**Supplementary table 6.** Predicted impact of a pre-existing antibody neutralization on viral kinetics using model M1.

| Antibody neutralization level at infection (ED<sub>50</sub>) | Median peak viral load (log<sub>10</sub> copies/mL) | Probability of peak viral load above the limit of detection | Probability of peak viral load above the threshold of infectivity |
| --- | --- | --- | --- |
| 0 | 6.6 | 100.0% | 73% |
| 10<sup>1</sup> | 6.4 | 100.0% | 66% |
| 10<sup>2</sup> | 6.2 | 99.9% | 59% |
| 10<sup>3</sup> | 6.1 | 99.9% | 54% |
| 10<sup>4</sup> | 5.9 | 99.2% | 46% |
| 10<sup>5</sup> | 5.7 | 98.6% | 41% |

